# An Updated Meta-analysis of Studies Comparing Conventional to Underwater Endoscopic Mucosal Resection for Colorectal Polyps

**DOI:** 10.1101/2022.05.17.22275225

**Authors:** Rahul Pamarthy, Hassam Ali, Syed Hamza Bin Waqar, Shiza Sarfraz

**Author notes:** Corresponding author: Hassam Ali, Department of Internal Medicine East Carolina University/Vidant Medical Center 600 Moye Blvd VMC MA Room 350 Mailstop #734 Greenville, NC 27834. **Statements and Declarations**. **Competing Interests** The authors have no relevant financial or non-financial interests to disclose. **Author contributions** HA: Conceptualization, Writing-Review, Supervision, Project Administration; RP: Writing-Original Draft, Writing-Review & Editing; SS: Writing-Original Draft, Writing-Review & Editing; SHBW: Writing-Original Draft, Writing-Review & Editing; EA: Investigation, Supervision, Editing, Proof reading. **Data Availability** The datasets generated during and analyzed during the current study are available from the corresponding author on reasonable request. **Ethics Approval** This is a meta-analysis. The University and Medical Center IRB at East Carolina University Research Ethics Committee has confirmed that no ethical approval is required. **Consent to participate:** Not applicable. **Consent for publication:** Not applicable.

## Abstract

**Background:** Underwater endoscopic mucosal resection (UEMR) is an emerging alternative to conventional endoscopic mucosal resection (CEMR). We intended to compare both techniques for colorectal polyp resection.

**Methods:** A comprehensive search of several databases to identify studies published until November 2021 was performed. Inclusion criteria included studies comparing UEMR to CEMR in adult patients. The calculation was done by standard meta-analysis methodology, and heterogeneity was assessed using the I2% statistics.

**Results:** 1029 polyps were resected with the CEMR technique and 1078 polyps with UEMR. UEMR was associated with an increase in the rate of overall en-bloc resection (Odds ratios (OR) 1.77; 95% CI, 1.42-2.22; P < .0001; I2 = 20%). Subgroup analysis showed an increase in the rates of en-bloc resection in polyps greater than 20 mm (OR 1.62; 95% CI, 1.17-2.25; P = 0.004; I2 = 33%). There was a reduction in the recurrence rate of polyps (P < 0.0001) in the UEMR cohort. Post-procedural bleeding or risk of perforation was not increased in either group. Resection times were shorter in UEMR (Mean difference, -8.09; P = 0.006).

**Conclusion:** UEMR is associated with lower recurrence rates and shorter procedure duration. In the future, UEMR may become the standard technique for colorectal polypectomy.

## Introduction

In 2020, colorectal cancer (CRC) was the second leading cause of cancer death in the United States and the fourth most common cancer by its incidence [1]. Data from the United States Surveillance, Epidemiology, and End Results (SEER) database suggest that CRC incidence is increasing in the under age 50 group and decreasing in older groups despite age being a significant risk factor for CRC [1]. Recently the guidelines have reduced the age of screening for CRC from 50 years to 45 years [2]. Endoscopic mucosal resection (EMR) has been the primary approach for large or sessile colorectal polyps [3].

The conventional EMR (CEMR) procedure utilizes a submucosal fluid injection underneath the polyp that helps create a cushion to separate the deeper muscularis mucosa from the superficial epithelial layer containing the lesion. This prevents the risk of perforation and electrical burn by increasing the gap between the current produced by electrocautery and the transmural space [4]. CEMR is a widely accepted and practiced technique among endoscopists and has replaced surgical resection almost completely. United States Multi-Society Task Force on Colorectal Cancer recommends using EMR for large (20 mm) non-pedunculated colorectal lesions [5]. However, it has been utilized for small, non-pedunculated colorectal polyps (4–9 mm in size) [6]. Literature shows that CEMR has been associated with high rates of incomplete resection and recurrence of follow-ups [3].

Binmoeller et al. first described the underwater endoscopic mucosal resection technique (UEMR) in 2012, in which the complete removal of large colorectal polyps was done without the need for submucosal injection. Removing intraluminal air in UEMR reduces colonic wall tension, permitting the colon wall to assume it’s natural collapsed state [7]. The safe technique demonstrated certain advantages over conventional EMR, resulting in further RCTs and cohort studies [8-13].

Multiple studies have been conducted comparing CEMR to UEMR and report good results in favor of UEMR and both in efficacy and safety [14,15]. Data comparing UEMR to CEMR has been systematically reviewed previously. However, the analysis were limited by the number of studies or inclusion criteria, including only larger polyps > 20mm [16,17]. Since those analysis, more trials have been published, adding strength to current literature regarding the safety and effectiveness of CEMR vs. UEMR for colorectal polyp resection [6,18].

## Methods

### Literature Search

We performed a comprehensive literature search across multiple databases from inception to November 2021. This included MEDLINE/PubMed, The Cochrane Library, Google Scholar, and Scopus.

Our inclusion criteria included studies that compared CEMR and UEMR for the resection of colorectal polyps irrespective of polyp size. The primary outcomes were en-bloc resection rates, and secondary outcomes were rates of polyp recurrence, resection time, post-procedure bleeding, and intra or post-procedural perforation. We did not include R0 or incomplete resection in our meta-analysis due to fewer trials reporting these. Our Search inquiries included keywords like Underwater, endoscopic mucosal resection, recurrence, colorectal polyp, colorectal lesion, and en-bloc resection. Additionally, the article bibliographies were reviewed to detect articles missed on the initial search. All studies with adequate data were included, irrespective of sample size, setting, polyps size, and geography. Studies done in patients with age less than 18 years and studies not published in the English language were excluded from our analysis. The flowsheet for study selection is given in Supplementary Fig 1.

### Data Extraction

Two reviewers (H.A., S.W.) reviewed the articles to extract data independently. Disagreements were settled by consensus among both authors.

### Study Quality Assessment

For randomized, controlled trials (RCTs), the Cochrane risk-of-bias tool was used to assess the quality of studies. It is a validated tool to detect bias in RCTs within areas like randomization, allocation concealment, selective reporting, blinding, and outcomes. Retrospective studies quality was assessed on the Newcastle-Ottawa scale (NOS). Selection of study groups, comparability of the groups, and ascertainment of either the exposure or outcome of interest for case-control or cohort studies, respectively, were evaluated. A study with a score >6 on the NOS was considered to be of good quality.

### Definitions and Outcomes

En-bloc resection was defined as resection of a lesion virtually without dissection in a single piece. Post-procedure bleeding was defined as bleeding (either early or late) occurring after the procedure. Recurrence was defined as adenoma recurrence up to 6 months after initial resection demonstrated on endoscopy or histology.

### Statistical Analysis

Dichotomous variables were consolidated to report odds ratios (OR) with 95% confidence interval (CI) and P-value. Continuous data were reported as mean differences (MD) for continuous data with 95% CI. Rates of primary and secondary outcomes were compared for polyps removed via CEMR to the UEMR. This meta-analysis utilized two models: The Mantel– Haenszel model and the DerSimonian and Laird model. The Mantel– Haenszel test (fixed-effect model) was used for outcomes with no heterogeneity. The DerSimonian and Laird (random-effects model) was used if significant heterogeneity was present. The I^2^ measure of heterogeneity was used to detect statistically significant heterogeneity. Values were classed as mild (I^2^ < 30%), moderate (I^2^ 30–50%), and considerable (I^2^ > 50%). The risk of publication bias was assessed using funnel plots. RevMan 5.3 (Review Manager, version 5.3; The Nordic Cochrane Centre, The Cochrane Collaboration, Copenhagen, Denmark) was utilized for statistical analysis. Funnel plots were investigated for the presence of publication bias. This review was initiated and summarized per the preferred reporting items for systematic review and meta-analysis (PRISMA) guidelines [19].

Out of 2107 polyps included in the primary outcome, 1078 polyps were resected with the CEMR technique and 1029 polyps with UEMR. Five included studies were randomized controlled trials; one was prospective, while the remaining five were retrospective cohort studies. Recurrence interval was reported in five studies and was up to six months. The baseline study characteristics of included studies are given in table 1.

**Table 1.** Characteristics of included studies

| Author | Year | Study design | No. of patients | No. of polyps | Inclusion Criteria | Polyp Criteria | Primary outcome | Recurrence Interval |
| --- | --- | --- | --- | --- | --- | --- | --- | --- |
| Liverant et al. [8] | 2016 | Retrospective | 67 | 67 | - | - | technical success of UEMR | 3–6 months |
| Cadoni et al. [9] | 2017 | Retrospective | 287 | 381 | - | - | number of en-bloc and R0 resections (any polyp morphology and size) | - |
| Schenk et al. [10] | 2017 | Retrospective | 99 | 135 | - | $\geq 15$ mm without prior attempted resection | Complete macroscopic resection | 3–6 months |
| Chien et al. [14] | 2019 | Retrospective | 242 | 242 | Age $\geq 20$ years | $\geq 10$ mm, nonpedunculated | En-bloc resection rate | - |
| Rodriguez et al. [15] | 2019 | Retrospective | - | 162 | | $\geq 15$ mm | Technical success | 3–6 months |
| Yamashina et al. [11] | 2019 | RCT | 210 | 210 | Age $\geq 20$ years | 10-20 mm, nonpedunculated | R0 resection rate (en-bloc resection + negative margins) | - |
| Mouchli et al. [13] | 2019 | Retrospective | 190 | 190 | - | - | recurrence rates and adverse events between UEMR and CEMR | - |
| Hamerski et al. [12] | 2020 | RCT | 303 | 303 | - | $\geq 15$ mm nonpedunculated, laterally spreading tumors, not located in appendiceal orifice, ileocecal | Curative resection rate (proportion with no endoscopic evidence | 3–6 months |

|  |  |  |  |  |  | valve, or<br>dentate line | of recurrence<br>at 3–6<br>month<br>follow-up) |  |
| --- | --- | --- | --- | --- | --- | --- | --- | --- |
| Yen et al. [23] | 2020 | RCT | - | 118 | Age ≥ 18 years | >5 mm, nonpedunculated without evidence of deep submucosal invasion | Incomplete resection rate (post resection positive margins) | 3–6 months |
| Zhang et al. [6] | 2020 | RCT | 130 | 142 | - | non-pedunculated colorectal polyps (4–9 mm in size) | Complete and en-bloc resection rate | - |
| Nagl et al. [18] | 2020 | RCT | 147 | 158 | Age ≥ 18 years | sessile or flat colorectal polyps between 20 and 40 mm | En-bloc resection rate | 6 months |

RCT: Randomized Controlled Trial, UEMR: Underwater Endoscopic Mucosal Resection, CEMR: Conventional Endoscopic Mucosal Resection

All six cohort studies had a NOS score > 6 indicating good quality. The cohorts were representative of the population. Follow-up intervals allowed adequate assessment of outcomes. There was no evidence of attrition bias skewing the data in these cohort studies. Two studies used the propensity match mechanism to control for differences in demographic characteristics among cases and controls. The remaining cohort studied did not report any significant differences in baseline characteristics of the population groups. The Cochrane risk of bias tool reported that none of the RCTs had a risk of attrition bias due to incomplete data. Additionally, In all five RCTs, endoscopists were not blinded to the intervention, which raised concern for outcome assessment bias. Outcomes for each study were prespecified, and post hoc analysis was not used to report data. NOS and the Risk of bias assessment for these studies can be seen in supplementary table 1 and supplementary Fig 2.

## Results

### Primary outcome (En-Bloc Resection)

Ten studies were used to analyze en-bloc resection and included 1917 polyps. 58.57% of polyps in CEMR group had en-bloc resection, while 68% in UEMR. Underwater EMR was associated with a significant increase upto 77% in the rate of en-bloc resection (OR 1.77; 95% CI, 1.42-2.22; P < 0.0001; I^2^ = 20 (Figure 1a and 1b). On subgroup analysis for polyps ≥20 mm, 34.3% had en-bloc resection in CEMR group vs. 43.2% in UEMR. UEMR was associated with significant increase upto 62% in the rates of en-bloc resection in polyps greater than 20 mm in size (OR 1.62; 95% CI, 1.17-2.25; P = 0.004; I^2^ = 33%) (Figure 2a and 2b).

**Figure 1.**
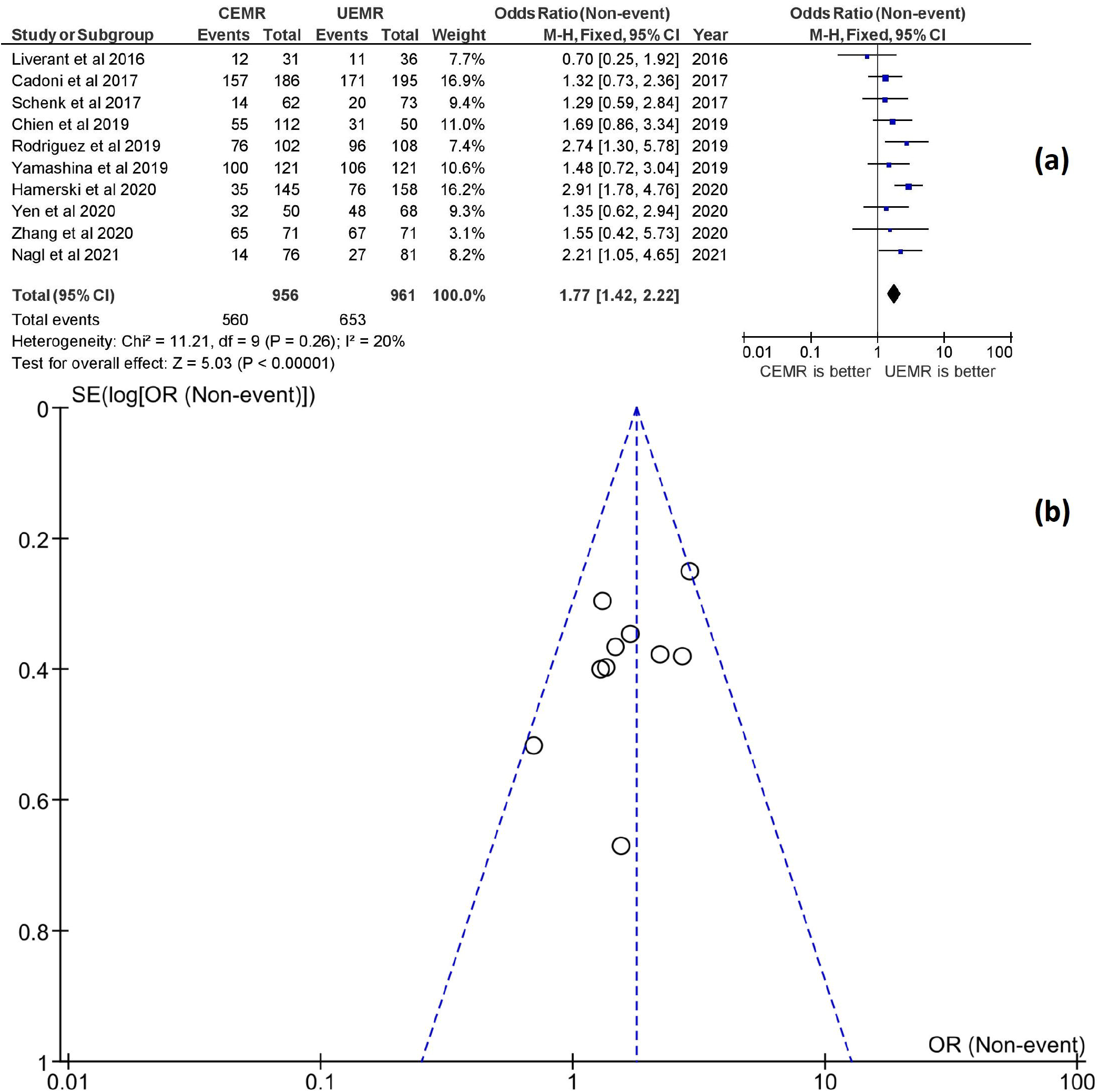
En-bloc resection rates in UEMR versus CEMR. a showing forest plots; b showing funnel plots

**Figure 2.**
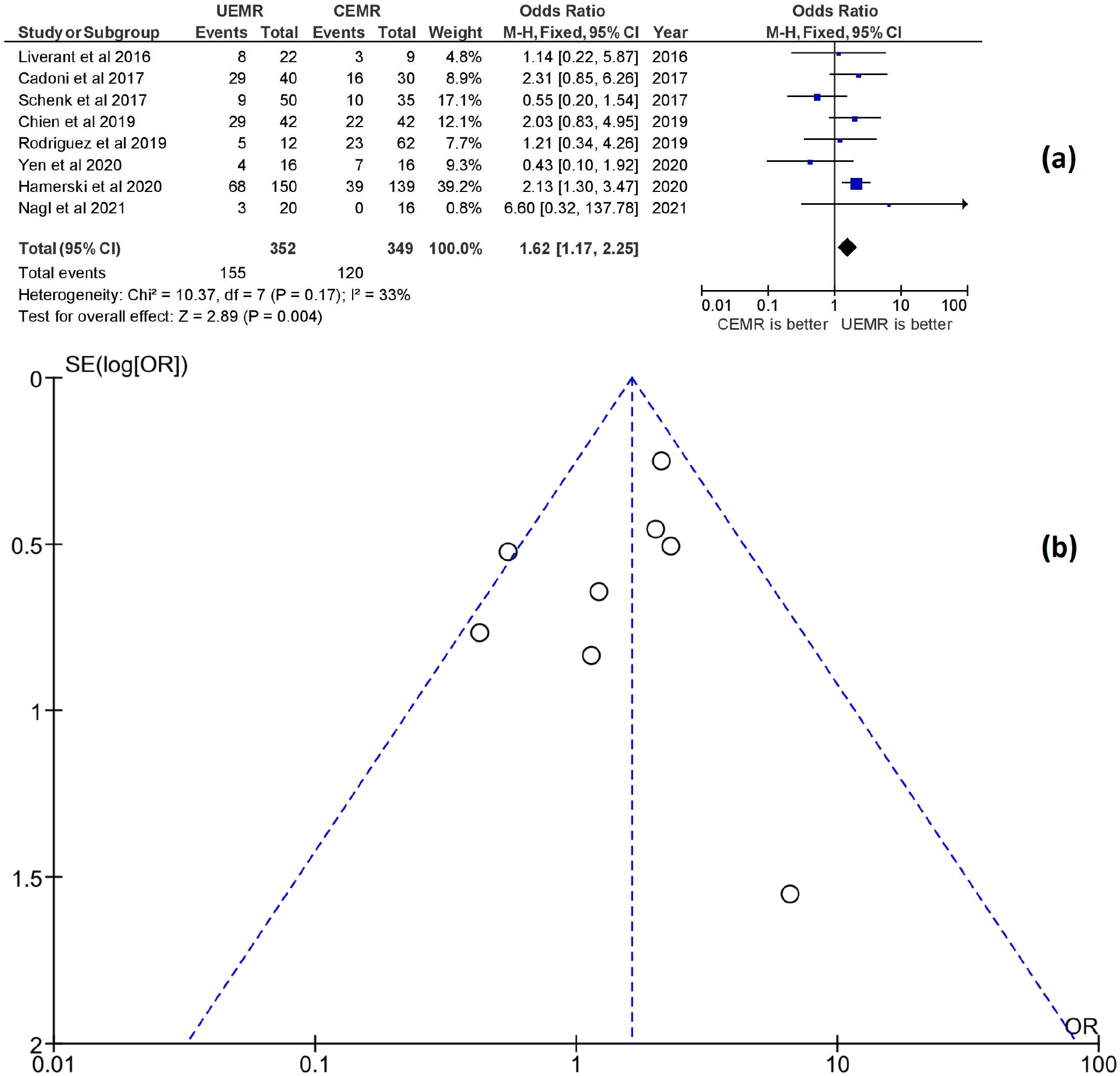
En-bloc resection rates in UEMR versus CEMR in polyps ≥20 mm. a showing forest plots; b showing funnel plots

### Secondary outcomes (Recurrence, Resection Time, postprocedural Bleeding, Perforation)

Six studies examined the recurrence rate and included 642 polyps. 3-6 months recurrence rate was 19.8% in the CEMR group compared to 7.6% in UEMR. UEMR was associated with a significant in the rate of recurrence (OR 0.33; 95% CI, 0.20-0.55; P < 0.0001; I^2^ = 0%) (Figure 3a and 3b). The recurrence rate based on polyp size was not calculated as it was not part of our outcomes.

**Figure 3.**
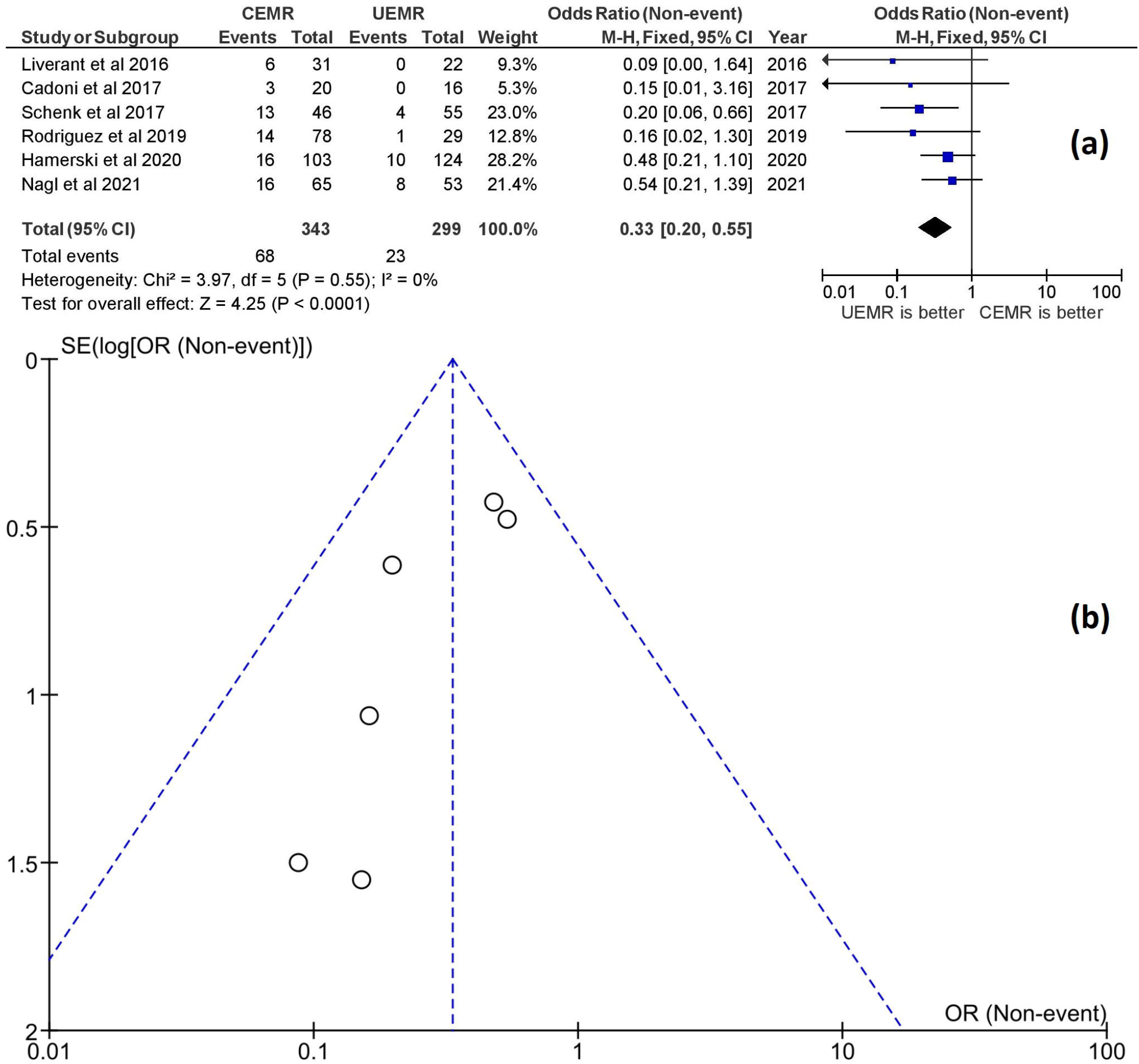
Forest plot of recurrence rates after UEMR versus CEMR at 3- to 6-month follow-up. a showing forest plots; b showing funnel plots

All eleven studies reported postprocedural bleeding and included 2107 polyps. Despite having higher power and low heterogeneity, there was no significant difference in the risk of early or late postprocedural bleeding between UEMR and CEMR (OR 0.89; 95% CI, 0.58-1.36; P = 0.59; I^2^ = 0%) (Figure 4a and 4b).

**Figure 4.**
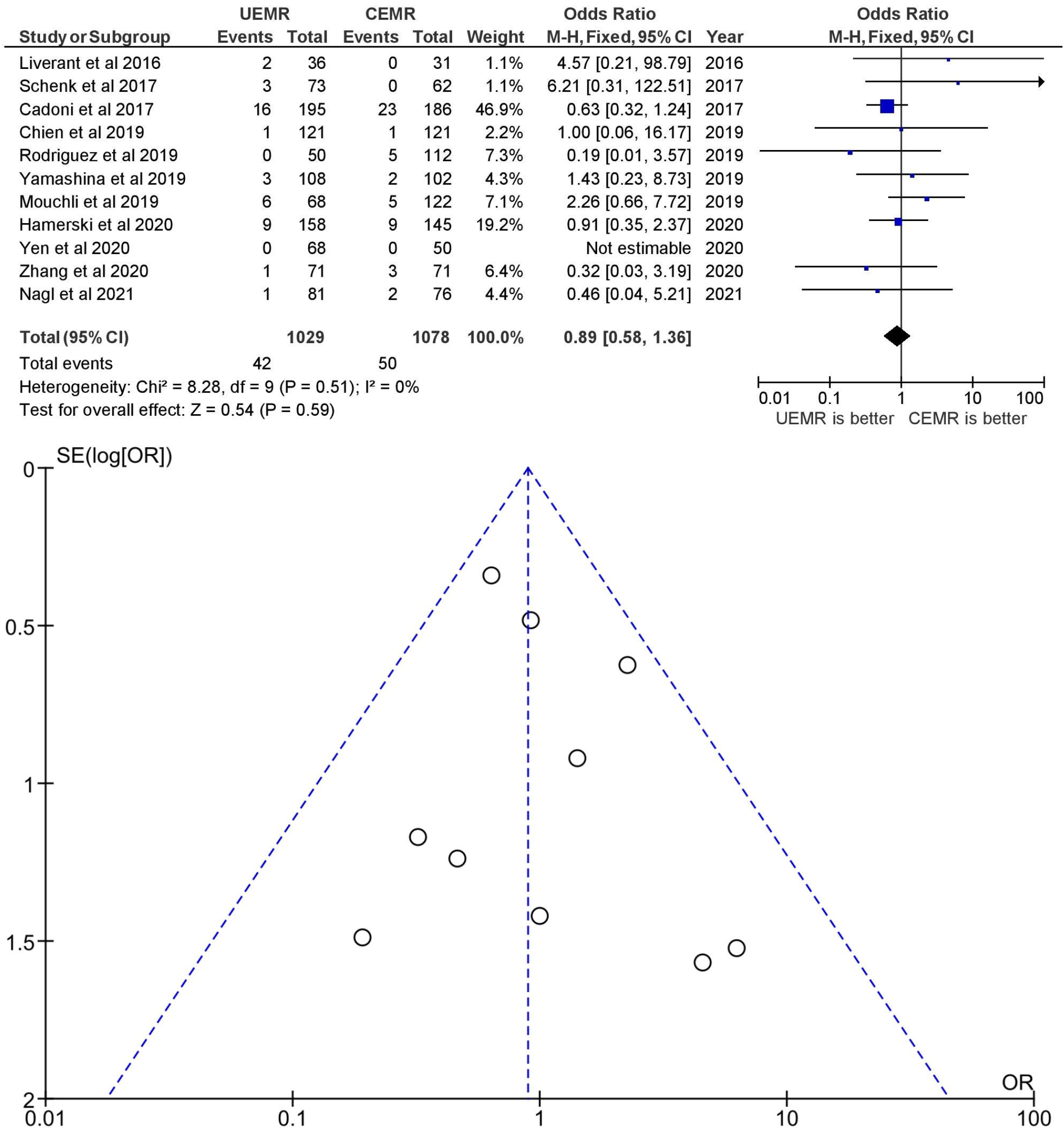
Forest plot of overall postprocedural bleeding rates in UEMR versus CEMR. a showing forest plots; b showing funnel plots

Ten studies reported perforation and included 1917 polyps in both groups. The risk of perforation was not significantly associated with CEMR or UEMR (OR, 0.73; 95% CI, 0.18-2.90; P = 0.66; I^2^ = 0%) (Figure 5a and 5b).

**Figure 5.**
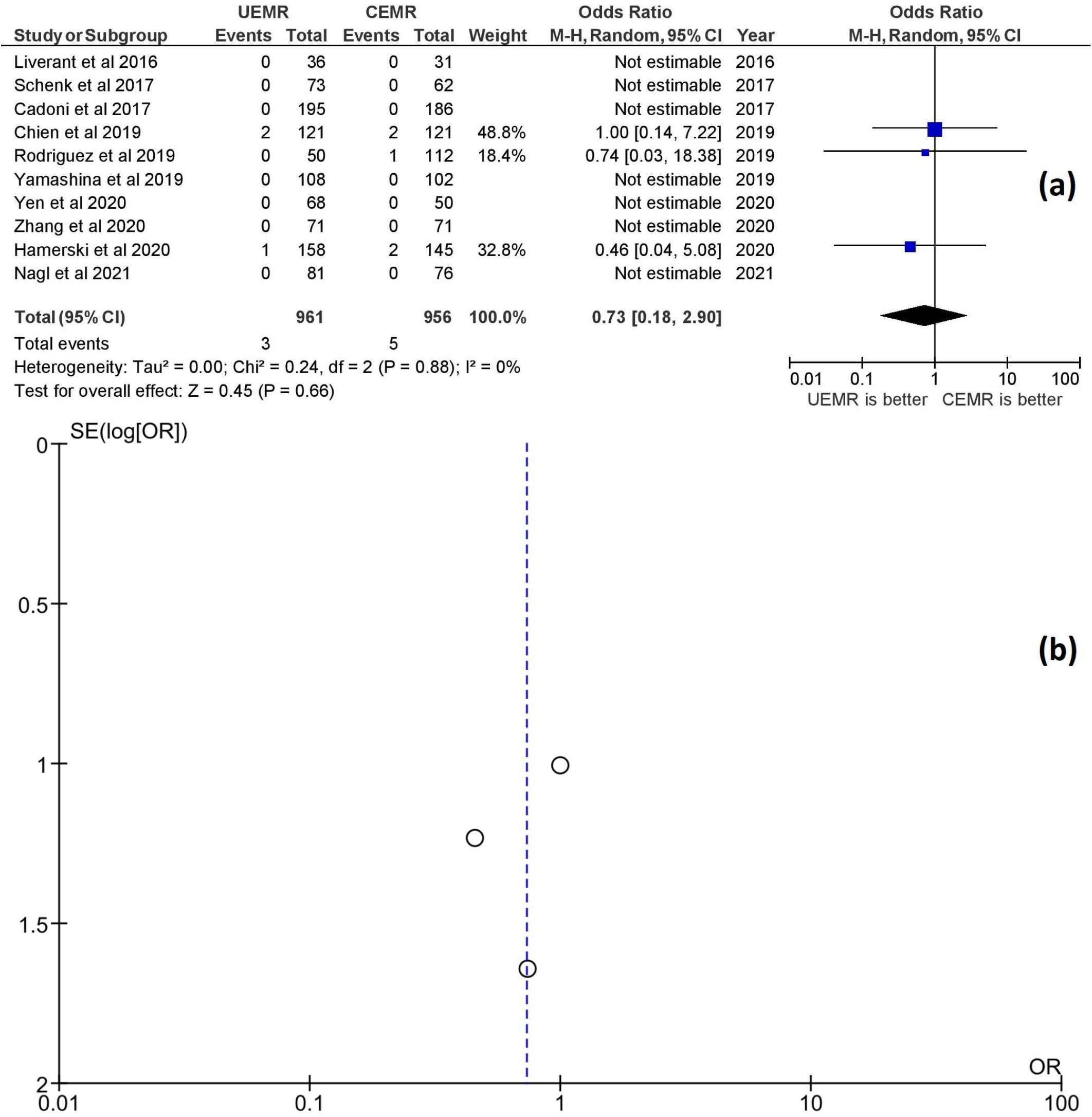
Forest plot of perforation rates in UEMR versus CEMR. a showing forest plots; b showing funnel plots

Four studies observed resection time and included 864 polyps. UEMR was associated with 8 minutes shorter procedure time than CEMR (Mean difference, -8.09; 95% CI, -13.82, -2.35; P = 0.006; I^2^ = 94%) (Figure 6a and 6b).

**Figure 6.**
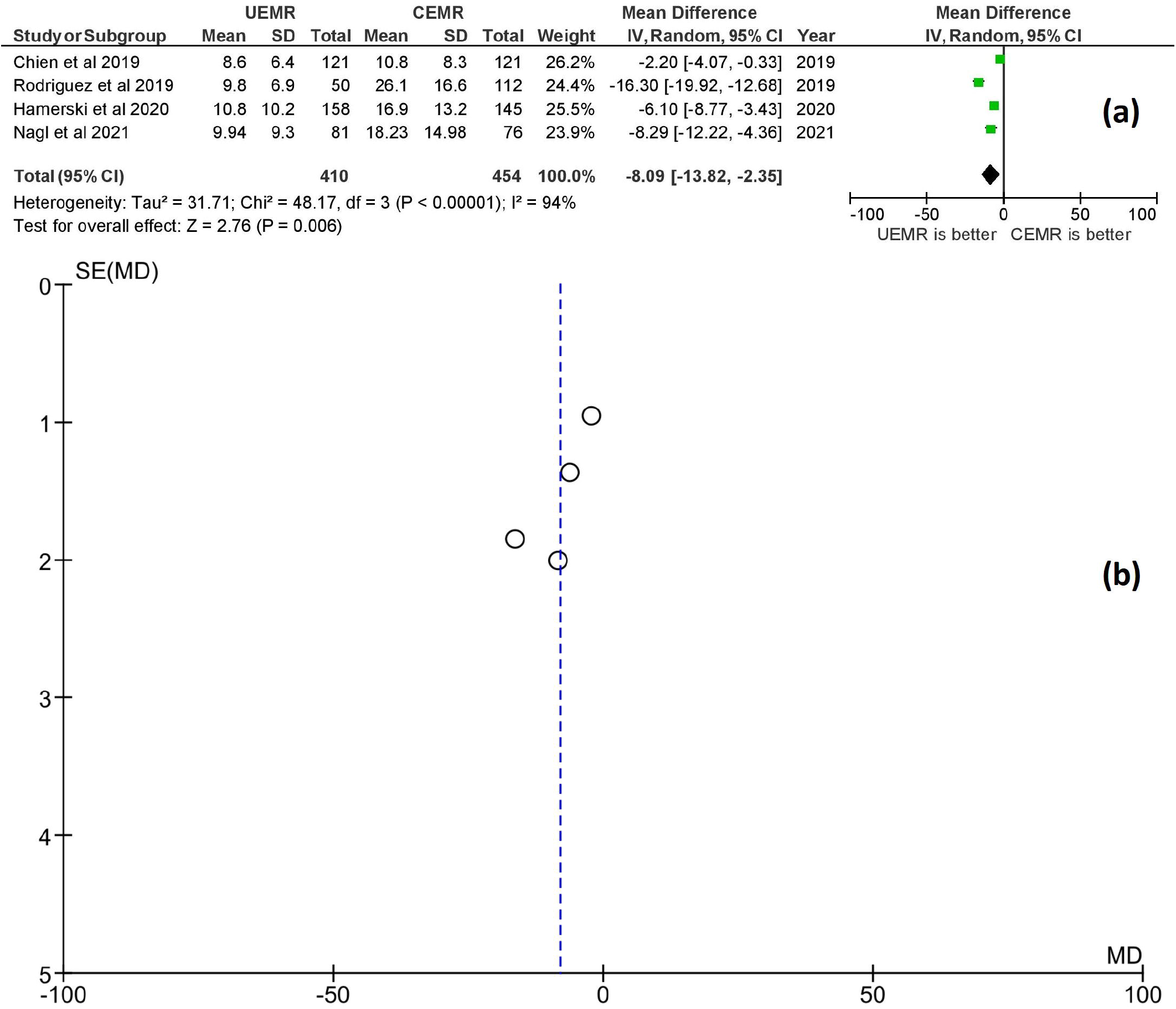
Forest plot of mean resection times for UEMR versus CEMR. a showing forest plots; b showing funnel plots

## Discussion

The current literature suggests that UEMR is efficacious and safe for the removal of colorectal polyps. The present study is the most recent systematic review and meta-analysis that includes recent trials and does not limit itself to the number of studies or polyp size [17, 20]. A total of eleven studies, including 2107 polyps, were included in this analysis: 1029 polyps were resected with CEMR and 1078 polyps with UEMR. The primary outcome proposes that UEMR could be the preferred method of colorectal poly resection for en-bloc removal for polyps as small as 5mm [6]. UEMR was also demonstrated to be better for polyps greater than 20mm on a sub-analysis. UEMR is associated with a lower recurrence rate up to 67 % per this analysis. The lower recurrence rate by UEMR could be secondary to higher en-bloc resection and lower piecemeal resection, which is associated with residual neoplasia [21].

The analysis show that UEMR can achieve increased en-bloc resection rates with no significant increase in side effects like the risk of perforation or post-procedural bleeding compared to EMR. Additionally, these side effects show a downward trend in patients who underwent UEMR compared to CEMR, which points towards a potential favorable safety profile of UEMR. The increased colonic wall tension secondary to gas insufflation in conventional EMR can flatten the polyp, complicating entrapment by snare and more residual tissue. This equated to lower en-bloc resection and higher recurrence. There is also a risk of submucosal microscopic seeding during submucosal injection in CEMR, contributing to a higher recurrence rate. There is a risk of microscopic seeding during submucosal injection in CEMR that can also contribute to a higher rate of recurrence [16,22]. UEMR provides better visualization due to a natural optical magnification/refraction effect underwater, enabling the gastroenterologist to have improved polyp detection. This leads to better en-bloc resections, resulting in lower recurrence rates and reduced procedure times [23]. Debris, blood, or dye can restrict vision in conventional EMR; however, the area of resection when underwater is kept clear, resulting in better visualization and inspection for polyps. Moreover, the wrinkling of colonic mucosa underwater provides for better friction with the snare and complete polypectomy. This could be the reason behind better en-bloc resection rates of larger polyps ≥20 mm seen in our analysis.

The reduced resection times with UEMR can also be explained by the less intensive preparation than CEMR, which involves determining the submucosal plane for injection, injection to optimize polyp positioning, and exchanging the needle for a snare, all of which consume more time. Moreover, the injection tends to expand the mucosal area of interest laterally and flatten over time, lengthening resection time. Due to lower resection times, gastroenterologists may abandon submucosal injections in the future and adopt UEMR ultimately. There was increased heterogeneity for analysis for resection time, possibly due to increased mean resection time in the CEMR cohort by Nagl et al. and Rodriguez et al. [15,18].

On pooled analysis, bowel perforation was less in UEMR (3 patients) than CEMR (5 patients). Perforation rates of CEMR have been described to range up to 1.5 % [24]. Pooled analysis in this study showed a perforation rate of 0.5% in the CEMR group and 0.3% in the UEMR group. Despite this, there was no statistically significant difference between both groups, although UEMR shows a better trend. The submucosal injection in CEMR helps to lift the polyp and avoids deep thermal burns. However, if the submucosal injection is misdirected, it can lead to perforation and muscle injury. This can be avoided with UEMR as the water submersion reduces colonic wall tension, and the submucosal fat moves away from muscularis propria resulting in a lower complication rate. Still, the perforation rate is lower among both groups, and UEMR has its benefits.

Post-procedural bleeding was 4% in UEMR compared to 4.6% in CEMR on pooled analysis without significant differences between the two groups. The submucosal injection needle could explain slightly more bleeding chances in CEMR than UEMR [25].

The strength of our study includes a systematic literature search, concise inclusion criteria, inclusion of studies with adequate data after precise evaluation of quality, and addition of studies missed on previous analysis due to search strategy or recent publications. Previous meta-analysis were limited either in the number of studies or polyp size [16,17]. Our analysis has its limitations, most of which are present with any meta-analysis. The studies included are not representative of a general population as they were performed in tertiary care centers at specific locations. Retrospective studies are weighted more and can contribute towards selection or confounding bias. The sample size for the timing of UEMR vs. CEMR was the smallest in our study. Nevertheless, this study is the most recent available literature that comprehensively compares the clinical outcomes of UEMR to CEMR for colorectal polyp resection and side effect profile.

## Conclusion

UEMR is an attractive alternative to CEMR with higher en-bloc resection rates, quicker resection time, and lower recurrences. The safety profile of both CEMR and UEMR is comparable, although UEMR did slightly better in each aspect. Endoscopists experienced in CEMR can quickly learn UEMR, and it may replace CEMR altogether in the future.

## Data Availability

All data produced in the present study are available upon reasonable request to the authors as this is a meta-analysis.

## Aknowledgment

None

## Notes

**Funding** The authors declare that no funds, grants, or other support were received during the preparation of this manuscript.

### Competing Interest Statement

The authors have declared no competing interest.

### Funding Statement

This study did not receive any funding

## References

1. H. Future incidence and mortality of colorectal carcinoma in the United States: an updated overview of risk factors and preventative measures. Explor Med. 2021;2:[Online First]. https://doi.org/10.37349/emed.2021.00063

2. Shaukat A, Kahi CJ, Burke CA, Rabeneck L, Sauer BG, Rex DK. Acg clinical guidelines: colorectal cancer screening 2021. Am J Gastroenterol. 2021;116(3):458–479. doi:10.14309/ajg.0000000000001122

3. Gaglia A, Sarkar S. Evaluation and long-term outcomes of the different modalities used in colonic endoscopic mucosal resection. Ann Gastroenterol. 2017;30(2):145–151. doi:10.20524/aog.2016.0104

4. Ferlitsch M, Moss A, Hassan C, et al. Colorectal polypectomy and endoscopic mucosal resection (Emr): european society of gastrointestinal endoscopy (Esge) clinical guideline. Endoscopy. 2017;49(3):270–297. doi:10.1055/s-0043-102569

5. Kaltenbach T, Anderson JC, Burke CA, et al. Endoscopic removal of colorectal lesions: recommendations by the us multi-society task force on colorectal cancer. Am J Gastroenterol. 2020;115(3):435–464. doi:10.14309/ajg.0000000000000555

6. Zhang Z, Xia Y, Cui H, et al. Underwater versus conventional endoscopic mucosal resection for small size non-pedunculated colorectal polyps: a randomized controlled trial : (UEMR vs. CEMR for small size non-pedunculated colorectal polyps). BMC Gastroenterol. 2020;20(1):311. doi:10.1186/s12876-020-01457-y

7. Binmoeller KF, Weilert F, Shah J, Bhat Y, Kane S. “Underwater” EMR without submucosal injection for large sessile colorectal polyps (With video). Gastrointest Endosc. 2012;75(5):1086–1091. doi:10.1016/j.gie.2011.12.022

8. Liverant ML, Yip B, Kwak N, et al. Su1690 Underwater Endoscopic Mucosal Resection (EMR) Shows a Higher Single Session Curative Resection Rate Than Conventional EMR Technique: A Single Center Experience [abstract]. Gastrointest Endosc. 2016;83:AB397.

9. Cadoni S, Liggi M, Gallittu P, et al. Underwater endoscopic colorectal polyp resection: Feasibility in everyday clinical practice. United European Gastroenterol J. 2018;6(3):454–462. doi:10.1177/2050640617733923

10. Schenck RJ, Jahann DA, Patrie JT, et al. Underwater endoscopic mucosal resection is associated with fewer recurrences and earlier curative resections compared to conventional endoscopic mucosal resection for large colorectal polyps. Surg Endosc. 2017;31(10):4174–4183. doi:10.1007/s00464-017-5474-4

11. Yamashina T, Uedo N, Akasaka T, et al. Comparison of Underwater vs Conventional Endoscopic Mucosal Resection of Intermediate-Size Colorectal Polyps. Gastroenterology. 2019;157(2):451-461.e2. doi:10.1053/j.gastro.2019.04.005

12. Hamerski C M, Wang A Y, Amato A et al. 121 Injection-Assisted versus underwater endoscopic mucosal resection without injection for the treatment of colorectal laterally spreading tumors: interim analysis of an international multicenter randomized controlled trial. Gastrointest Endosc. 2018;87:AB55–AB56.

13. Mouchli M, Walsh C, Reddy S et al. Sa1727 – Outcomes of gi polyps resected using underwater endoscopic mucosal resection (UEMR) Compared to conventional EMR (CEMR) Gastroenterology. 2019;156:S379–S380.

14. Chien HC, Uedo N, Hsieh PH. Comparison of underwater and conventional endoscopic mucosal resection for removing sessile colorectal polyps: a propensity-score matched cohort study. Endosc Int Open. 2019;7(11):E1528–E1536. doi:10.1055/a-1007-1578

15. Rodríguez Sánchez J, Uchima Koecklin H, González López L, et al. Short and long-term outcomes of underwater EMR compared to the traditional procedure in the real clinical practice. Rev Esp Enferm Dig. 2019;111(7):543–549. doi:10.17235/reed.2019.6009/2018

16. Garg R, Singh A, Mohan BP, Mankaney G, Regueiro M, Chahal P. Underwater versus conventional endoscopic mucosal resection for colorectal lesions: a systematic review and meta-analysis. Endosc Int Open. 2020;8(12):E1884–E1894. doi:10.1055/a-1287-9621

17. Choi AY, Moosvi Z, Shah S, et al. Underwater versus conventional EMR for colorectal polyps: systematic review and meta-analysis. Gastrointest Endosc. 2021;93(2):378–389. doi:10.1016/j.gie.2020.10.009

18. Nagl S, Ebigbo A, Goelder SK, et al. Underwater vs conventional endoscopic mucosal resection of large sessile or flat colorectal polyps: a prospective randomized controlled trial. Gastroenterology. 2021;161(5):1460-1474.e1. doi:10.1053/j.gastro.2021.07.044

19. Page MJ, McKenzie JE, Bossuyt PM, et al. The PRISMA 2020 statement: an updated guideline for reporting systematic reviews. BMJ. 2021;372:71. doi:10.1136/bmj.n71

20. Chandan S, Khan SR, Kumar A, et al. Efficacy and histologic accuracy of underwater versus conventional endoscopic mucosal resection for large (>20 mm) colorectal polyps: a comparative review and meta-analysis. Gastrointest Endosc. 2021;94(3):471-482.e9. doi:10.1016/j.gie.2020.12.034

21. Woodward TA, Heckman MG, Cleveland P, De Melo S, Raimondo M, Wallace M. Predictors of complete endoscopic mucosal resection of flat and depressed gastrointestinal neoplasia of the colon. Am J Gastroenterol. 2012;107(5):650–654. doi:10.1038/ajg.2011.473

22. Silva MA, Hegab B, Hyde C, Guo B, Buckels J a. C, Mirza DF. Needle track seeding following biopsy of liver lesions in the diagnosis of hepatocellular cancer: a systematic review and meta-analysis. Gut. 2008;57(11):1592–1596. doi:10.1136/gut.2008.149062

23. Yen AW, Leung JW, Wilson MD, Leung FW. Underwater versus conventional endoscopic resection of nondiminutive non-pedunculated colorectal lesions: a prospective randomized controlled trial (With video). Gastrointest Endosc. 2020;91(3):643-654.e2. doi:10.1016/j.gie.2019.09.039

24. Ponugoti PL, Rex DK. Perforation during underwater EMR. Gastrointest Endosc. 2016;84(3):543–544. doi:10.1016/j.gie.2016.01.021

25. Nett A, Binmoeller K. Underwater endoscopic mucosal resection. Gastrointest Endosc Clin N Am. 2019;29(4):659–673. doi:10.1016/j.giec.2019.05.004

