## Supplementary Fig 1 for "An Updated Meta-analysis of Studies Comparing Conventional to Underwater Endoscopic Mucosal Resection for Colorectal Polyps"

**Supplementary Fig. 1 : PRISMA flowsheet for data selection**

**Page MJ, McKenzie JE, Bossuyt PM, Boutron I, Hoffmann TC, Mulrow CD, et al. The PRISMA 2020 statement: an updated guideline for reporting systematic reviews. BMJ 2021;372:n71. doi: 10.1136/bmj.n71**

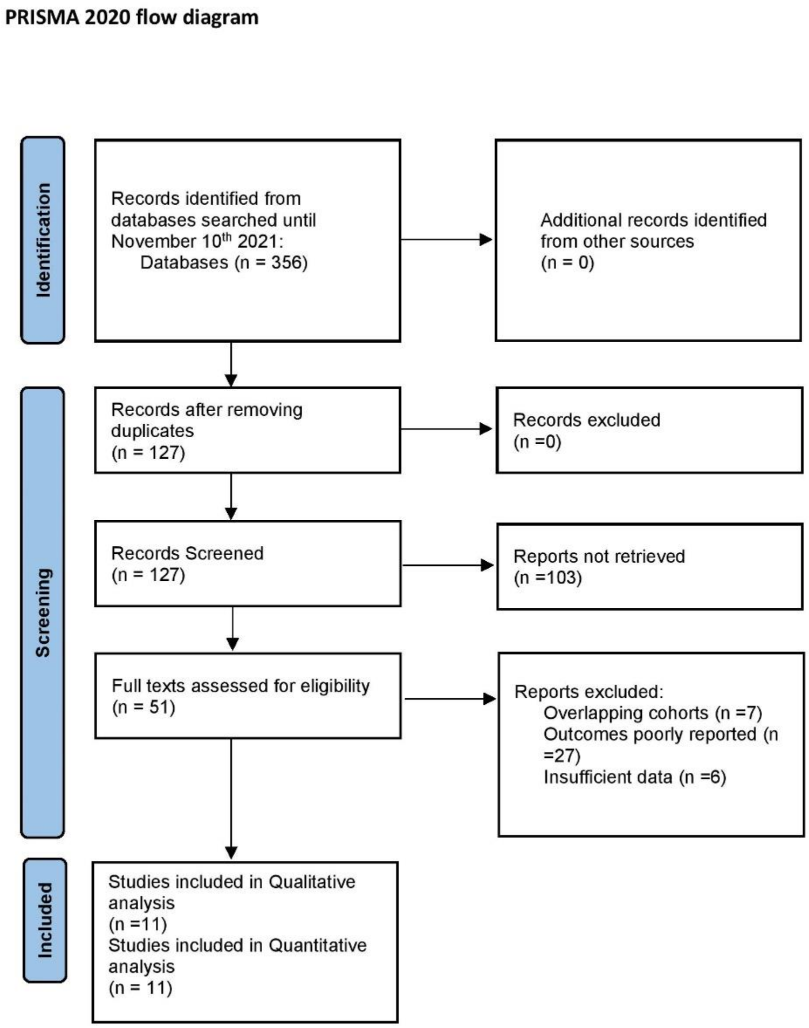

**Supplementary Table 1. Newcastle-ottawa scale (NOS) table summarizing the non-randomized studies included in our analysis.**

**Supplementary Table 1.** NOS for the risk of bias and quality assessment of NRSs

| Author | Year | Selection | | | Comparability | | Exposure | | | Total score |
|---|---|---|---|---|---|---|---|---|---|---|
| | | Adequate definition of patient cases | Representative ness of patient cases | Selection of controls | Definition of controls | Control for important or additional factors | Ascertainment of exposure | Same method of ascertainment for participants | Nonresponse rate | |
| Liverant et al. | 2016 | ⋆ | ⋆ | | ⋆ | ⋆ | ⋆ | ⋆ | | 6 |
| Cadoni et al. | 2017 | ⋆ | ⋆ | | ⋆ | ⋆⋆ | ⋆ | ⋆ | | 7 |
| Chien et al. | 2017 | ⋆ | ⋆ | | ⋆ | ⋆⋆ | ⋆ | ⋆ | | 7 |
| Schenck et al. | 2017 | ⋆ | ⋆ | | ⋆ | ⋆⋆ | ⋆ | ⋆ | | 7 |
| Rodriquez et al. | 2019 | ⋆ | ⋆ | | ⋆ | ⋆⋆ | ⋆ | ⋆ | | 7 |
| Mouchli et al. | 2019 | ⋆ | ⋆ | | ⋆ | ⋆⋆ | ⋆ | ⋆ | | 7 |

NOS, Newcastle-Ottawa scale; NRS, non-randomized study.

**Supplementary Fig 2. A revised Cochrane risk-of-bias tool for randomized trials included in our analysis.**

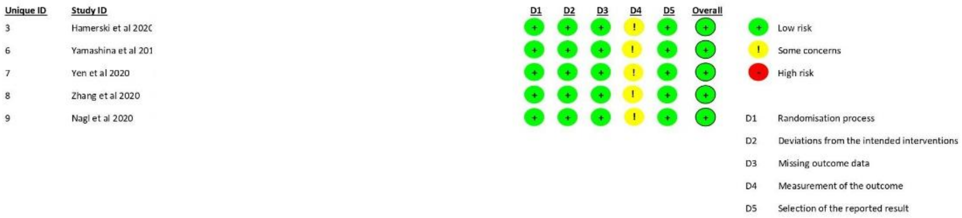
